# Adding a reaction-restoration type transmission rate dynamic law to the basic SEIR COVID-19 model

**DOI:** 10.1101/2021.07.13.21260408

**Authors:** F. Córdova-Lepe, K. Vogt-Geisse

## Abstract

The classical SEIR model, being an autonomous system of differential equations, has important limitations when representing a pandemic situation. Particularly, the geometric unimodal shape of the epidemic curve is not what is generally observed. This work introduces the *β*SEIR model, which adds to the classical SEIR model a differential law to model the variation in the transmission rate. It considers two opposite thrives generally found in a population: first, reaction to disease presence that may be linked to mitigation strategies, which tends to decrease transmission, and second, the urge to return to normal conditions that pulls to restore the initial value of the transmission rate. Our results open a wide spectrum of dynamic variabilities in the curve of new infected, which are justified by reaction and restoration thrives that affect disease transmission over time. Some of these dynamics have been observed in the existing COVID-19 disease data. In particular and to further exemplify the potential the model proposed in this article, we show its capability of capturing the evolution of the number of new confirmed cases of Chile and Italy for several months after epidemic onset, while incorporating a reaction to disease presence with decreasing adherence to mitigation strategies, as well as a seasonal effect on the restoration of the initial transmissibility conditions.

## 1 Introduction

A novel Coronavirus (SARS-CoV-2) emerged from the city Wuhan in China in December 2019 and has caused a devastating public health impact across the world [1]. As of June 28, 2021, COVID-19 has caused over 180 million confirmed cases and over 3.5 million deaths worldwide [2]. The curves for daily confirmed new cases of COVID-19 in different countries present a high variability in their geometric forms. Every such curve shows a sequence of outbreaks and valleys when observed over time, while the sharpness of the outbreaks and the length of the valleys can vary [3].

In fact, Fig. 1 shows the daily number of confirmed new cases (7 day moving average) from March, 2020, to January, 2021, for different countries. Most of them have experienced a second wave and others show even a third [3]. Some European countries show a sharp first outbreak followed by a plateau of low height (that lasted several months) before the second wave, whose peak out-measures the peak of the first outbreak by at least three-fold (see plots (a-b-c) in Fig. 1). On the contrary, several South American countries presented an initial exponential phase of several months, soon after which a second wave of similar peak height as the first occurs (see (d-e-f) Fig. 1). Finally, there are countries in which the curve of new cases of COVID-19 has shown extreme behaviors in some part of its evolution as compared to European and South American countries. For instance, Czech Republic has experienced a very weak first outbreak followed by a low plateau lasting for months; Iran did have a more pronounced first outbreak, but it was followed by a plateau of important height; and Indonesia experienced a less distinct first outbreak that resembles exponential growth when zoomed out (see respectively (g-h-i) in Fig. 1).

**Fig. 1:**
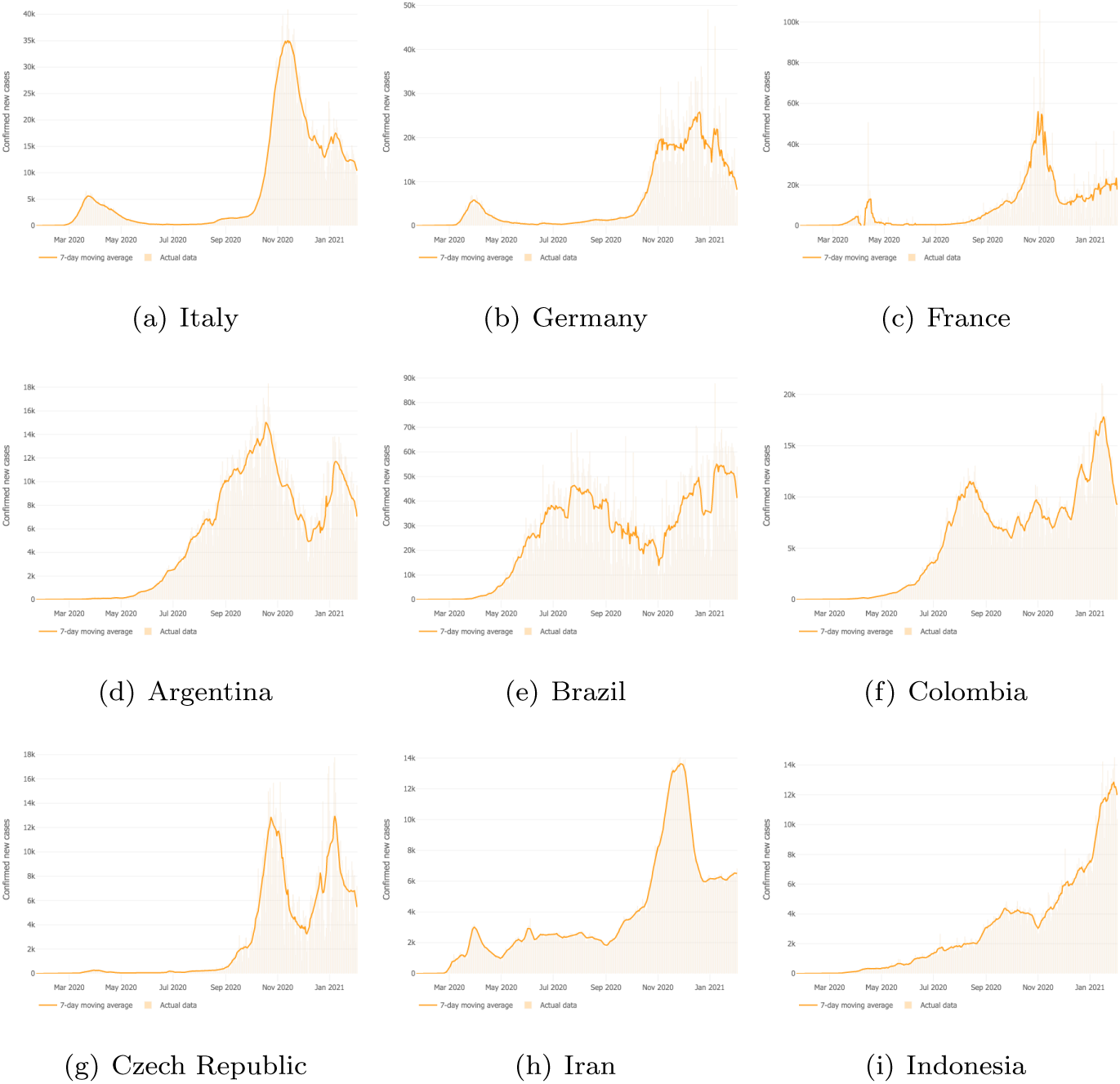
Confirmed new cases (7 day moving average) from March 2020 to January 2021. European countries ((a), (b) & (c)) showing a sharp first outbreak followed by a low height plateau (compared to the waves) lasting for several months, after which a second wave occurs of peak at least three times higher than the peak of the first wave. South American countries ((d), (e) & (f)) showing an initial exponential phase of several months that define an important first outbreak followed by a reduction in cases of a few months, after which a second wave as large as the first occurs. Other countries that show curves of new cases that reflect extreme situations as compared to European and in South America ones. Czech Republic (g) shows a very weak first outbreak followed by a long plateau. Iran (h) pictures a plateau of important height after the first outbreak. Indonesia (i) shows a less distinct first outbreak that shapes an exponential behavior when zoomed out. Plots downloaded from “ *Maps & Trends. New cases of COVID-19 in world countries*” [3].

Classical compartmental models based on the classical Kermack & McKendrick SIR model [4] with constant parameters often used to model epidemics do not reflect the behavior over several months described above. Neither the (*β, γ*)SEIR model for a population of size *N* – which is compartmentalized into susceptible (*S*), exposed (*E*), infectious (*I*) and removed (*R*)– given by

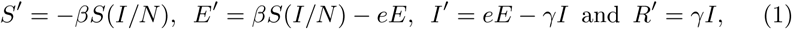

nor extensions of it have been efficient in adjusting the data well beyond the first epidemic outbreak when considering the transmission rate *β* and the removal rate *γ* constant. This is due to the unimodality of the active-infected-curve those models provide, i.e. one bell-shaped infection curve and an epidemic growth limited by the proportion of susceptible individuals [5, 6].

In general, the epidemiological data series do not reflect that the percentage variation of susceptibles per proportion of active cases, i.e. |*S′/S*|*/*(*I/N*), is approximately constant, as is assumed in the aforementioned classical epidemiological models. In fact, there exists literature that evidences the changing temporal behavior of disease transmission in epidemic or pandemic situations [7, 8,9, 10, 11, 12, 13, 14,15]. In particular, there are studies using mathematical models– some aiming to understand COVID-19 transmission– that include the decrease in the transmission rate [7, 16, 9,17,18, 19, 20, 21], and some incorporating human behavioral factors as part of the cause for a temporal change in transmission. For instance, in [7] the behavior of the transmission rate– providing exponential saturation for a large number of infectives– for three consecutive months is shown for four geographical settings: worldwide, United States, Russia and Canada. For each setting, the trend is a decrease in the transmission rate for then stabilizing at a value several times lower than initially. They as well include a parameter representing mask wearing within their transmission rate. Finally, this study also shows that the recovery rate *R′/I* is for each setting much more stable than the transmission rate. Supporting the idea that the lack of efficiency for the classical compartmental models to adjust well to data is due to that the transmission rate *β* is assumed constant over time. We present in more detail examples found in the literature about non-constant transmission rates within compartmental models in Sec. 2.

In this article we attempt to break the unimodality of the active-infected-curve of the classical epidemiological models. We introduce a novel way to model the behavior of the transmission rate *β*, considering a balance equation between a reaction rate and a restoration rate; and including the resulting dynamic law for the transmission rate into the classical SEIR model. The paper is structured in the following way: In Sec. 2 we provide some understanding about the transmission rate of infectious diseases. In Sec. 3 we introduce and describe a new basic model, which we call *β* SEIR model, by adding to the classical SEIR the aforementioned dynamic law for the transmission rate, and show some mathematical analysis. In Sec. 4 we provide numerical results and finally in Sec. 5 the discussion and conclusions.

## 2 The transmission rate

There exist at least two groups of epidemic control measures. The first, aims to reduce the population that is being hit by the disease, i.e., the susceptible population. Such measures are, for example, vaccination or limiting the mobility of individuals. The second, intents to reduce the *force of infection* that is defined as the product of three quantities: number of close contacts per unit of time of a susceptible individual (*p*_*C*_), probability that a close contact is with an infectious individual (*p*_*I*_) and probability of transmission given a close contact with an infectious (*p*_*T*_). Reducing *p*_*I*_ results in that per unit of time, there exist fewer active cases in the population, which is accomplished by eradication, i.e., removal from the system (e.g., slaughtering of sick animals which is widely used in animal epidemics, or banishing infectious individuals as was done aforetime), or by applying actions for a rapid recovery. Notice that the product of *p*_*C*_ and *p*_*T*_ is called the *transmission rate* and is usually denoted by *β* (see e.g. Eqn. (1)). Hence, the objective of most mitigation strategies that aim to reduce the force of infection, aim to reduce the transmission rate (lower *β*) by either increasing physical distance and hence reducing the number of close contacts (lower *p*_*C*_) or blocking the transfer of pathogens to a new host (lower *p*_*T*_). There are secondary measures such as for example reducing population movement (which is not reducing physical distancing nor blocking transmission), which make close encounters less likely.

When a highly transmissible disease with high mortality or morbidity invades a population of mostly susceptible individuals, and a vaccine is not in sight in the short term (as was initially the case for COVID-19), health authorities’ only way for reducing morbidity and mortality is mitigation, while the general population’s duty is to comply to the new norms and desired behavior. In other words, the efforts are put into reducing the transmission rate *β*. In this sense, *β* is a time-spacial dependent variable, i.e. it changes according to time and location. Also less controllable physical-environmental aspects in relation with the bio-chemical characteristics of the pathogen may influence it (but we consider those factors constants in this study). It is also worth mentioning that in general populations, individuals may live and participate in several cultural regions, which may also determine the variability of the transmission rate. We will assume in this study that the population stays within its territory during the time horizon studied, behaving homogeneously in this respect. We suppose that the disease studied will have a base line transmission rate *β*_0_, that we will call *natural transmission rate*, measured for a population that is initially free from the disease and does not consider any mitigation strategies or personal protective measures.

One of the characteristics of COVID-19 was that it has had a large media coverage since the first confirmed cases appeared in December 2019. This provoked sentiments of fear in the general population and played an educational role for pandemic preparedness (e.g. global media emphasizing on washing hands and physical distancing) before the imminent arrival of COVID-19 in many countries. It shaped how countries would confront COVID-19 right from the appearance of their first confirmed case and even before. The reproduction numbers corresponding to different geographical locations was most likely to be between 2 and 5 [22, 23], and was shown to rapidly decrease during the first weeks of the pandemic [13, 12, 14], however reaching values above one that nevertheless allowed COVID-19 expansion.

To consider this decreasing effect, many authors assume an exponential decay of the transmission rate for a certain amount of time, for example, in [18, 19] they assumed in their continuous time model *β*(*t*) = *β*_0_ exp(*−b*_0_*t*), *t ≥*0, or in in [20, 21] they defined *β*_*k*_ = *β*_0_ *a*^*k*^, 0 *< a <* 1, *k ≥*0, where *k* is a day-counting integer in their discrete time model. In order to extend the horizon of validity of their model some authors consider an exponential decrease from *β*_0_ to a minimum positive bound [24, 25]. To understand the rate of decrease from that baseline natural *β*_0_ and identify a time varying *β*(*t*) transmission rate, some researchers use mathematical expressions and the data on active cases *I*(*t*) and removed cases *R*(*t*) in a population of constant size *N*; for instance, one can use *β*(*t*) = *−NS′/*(*SI*) as in [7], which can be obtained from a SIR model and approximating the derivatives using the finite differences on one week running averages; or one could use *β*(*t*) = *γ* + *I′/I* at the beginning of an outbreak, assuming *S ∼N* in the SIR model.

More time-varying transmission rates have been considered within mathematical models. For instance, the authors in [9] capture the early decreasing trend of COVID-19 in Malaysia using a time varying exponential decay log function *β*(*t*) = *zβ*(1 *− p*)^*t*^ for the transmission rate in an SIR model, that uses a fractional term *z* to measure the effectiveness of interventions and a proportion *p* to account for depletion. In the literature there are studies that, in order to capture realistic disease transmission, assume non-linear functions of *S* and *I* governing the force of transmission, as for instance in [16], where the force of transmission they use in an SIR-type model depends on the product of fractional powers of *S* and *I*. They use the model to fit COVID-19 data of Italy, Germany, France and Spain. In [26] the authors include in an SIR model time-varying transmission rate, assuming that the probability of transmission of a susceptible is *βλ*_*t*_(*I*_*t*_*/N*), where *λ*_*t*_(*·*) is a random variable, they refer to this model as a Spatial-SIR model. In [27] the authors assume a contagion rate as a sum of a base-line transmission rate and a component that satisfies a first order linear differential equation to represent the effect of non-pharmaceutical interventions (NPIs).

Behavioral factors represented in the transmission rate are also considered by more authors in order to represent the changing dynamics of the transmission rate. For instance in [28] the authors study a model that incorporates a non-constant transmission rate *β* (*M*) that depends not only on the current number of infectious individuals but on *M*, representing an information index that summarizes the current and past history of disease prevalence. Part of their results discuss that social behavioral change may trigger oscillations. The study in [29] extends an SIR type model defining a transmission rate that captures the impact of school and workplace closure through a function of time. The changing behavior of this function is based on Imitation Dynamics [30] and describes population-level support dynamics for closure. The article in [31] also uses Imitation Dynamics and studies a population in which individuals develop and learn a behavior of mutual protection.

The novelty of this article is that at each time *t*, we consider the variation of the transmission rate between the end-points of the time interval [*t, t* + *Δt*] to be given according to a balance equation between two opposite thrives: a *reaction rate* (fraction of reduction per unit of time) and a *restoration rate* (fraction of increment per unit of time). It takes the form

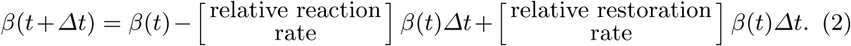

In what follows we are going to justify the functional forms of the reaction and restoration rates, as well as present the *β* SEIR model that incorporates the dynamic law for *β*. Further, we are going to analyze its effect on the shape of the main epidemiological curves.

## 3 The *β* SEIR Mathematical Model

In this section we present the dynamic law of the transmission rate *β* in order to introduce the *β* SEIR model and some mathematical analysis.

### 3.1 Transmission rate *β*

In this subsection we derive the form of the transmission rate at which susceptible individuals become exposed upon contacts with infectious. The only infectious class of the model is the *I* class. We model the case of an infectious disease transmitted directly from person to person, and assume that at the beginning of an outbreak, the appearance of first cases do not provide a reason for alarm and panic. Hence, initially the disease propagation is due to a high *natural-transmission rate* intrinsic to the population while no interventions to mitigate disease spread are in place. We call this *natural-transmission rate β*_0_. In general, *β*_0_ makes the disease expand rapidly, producing a fast initial increase in new cases.

We present a new form for the transmission rate, which is represented by a dynamic, time-dependent quantity that is governed by a balance equation between a *reaction rate, g*[*t, I*], and a *restoration rate, f* [*t, β*], i.e. the proportion by which the transmission rate decreases and increases per day, respectively, represented by

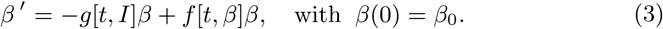

Next we justify the introduction of a reaction rate and a restoration rate and propose a functional form for each.

#### 3.1.1 Reaction rate

During severe epidemic outbreaks that attract huge public attention and media coverage due to for instance a high morbidity and/or mortality in the population, a steady increase in the implementation of measures that aim to reduce the transmission rate can be observed. As long as there is no licensed vaccine or treatment, these measures are mainly based on non-pharmaceutical interventions. Here we are interested in those directed to diminish the factors *p*_*C*_ and *p*_*T*_, whose product defines *β*, such as social distancing measures (that reduce the number of close contacts between people: large-scale or home quarantines, workplace nonattendance, travel restrictions, prohibition of social gatherings, school closures, etc.) that reduce *p*_*C*_, or blocking measures (that, given e contact, reduce the pass of the pathogen: hand-washing, respiratory etiquette, face-masks usage, etc.) that reduce *p*_*T*_, see [32, 33]. When fear governs the population, people react, complying with mandatory measures or adopting self-protecting measures to avoid infection [34]; we note that risk communication is also a factor to support the general public response [35]. It is to expect that the higher the severity of the disease is, the more effort is put into mitigation. In this sense, individuals’ reactions produce a decrease in the transmission rate from its initial *natural-transmission rate* value, *β*_0_. Hence, we define a *reaction rate* that we denote *g*[*t, I*], which is non-negative and positively correlated with the number of active cases *I*(*t*), in a way that it increases when *I*(*t*) does. It may also depend on other circumstantial conditions of the moment relative to the population. These we will discuss further later in the text.

We assume in this article, that the *reaction rate*, follows the Michaelis-Menten model [36] describing the reaction to the presence of the infectious (active cases) *I*(*t*) at any moment in time, i.e. we define

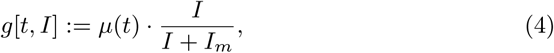

where we call *µ*(*t*) the reaction coefficient, which is a non-negative function that represents the daily maximum possible reduction at time *t*, and *I*_*m*_ *>* 0 is the half-saturation constant, i.e. is the number of active cases, where the reduction is half-maximal. Notice that the parameter *I*_*m*_ characterizes the population, i.e. it determines its sensibility to react to active cases.

#### 3.1.2 Restoration rate

It is important to observe that upon the appearance of a reaction rate there exist socio-environmental factors that tend to restore the transmission rate to the level observed at the beginning of the pandemic, i.e. to *β*_0_ [29, 37]. When in a certain location the health authorities cease to impose protective measures, e.g. the use of face masks, and individuals lost their initial fear, then the transmission rate that had been reduced due to these measures no longer stays low and returns to its natural level. Therefore, we introduce a *restoration rate* that at each instant *t, t >* 0, is responsible for an increase in the transmission rate. The *restoration rate* that we denote by *f* [*t, β*], is a non-negative function that correlates directly with the distance between *β*(*t*) and its natural value *β*_0_, as presented in the following equation:

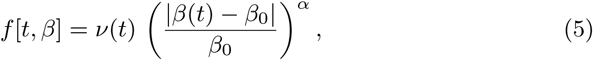

where *α ∈*ℝ is a positive exponent. We call *ν*(*t*) the restoration coefficient, which is a non-negative function that regulates the daily form of the restoration rate. Also, *f* is an increasing function of| *β*(*t*) *− β*_0_ |, and *f* [*t, β*_0_] = 0, which means that if the transmission rate *β*(*t*) reaches at a certain time point *t* its natural value, then there is no deviation to restore.

### 3.2 The *β* SEIR model: formulation and analysis

We incorporate the differential equation (3) to the classical SEIR model (1) obtaining the *β* SEIR model, given by the following system of equations

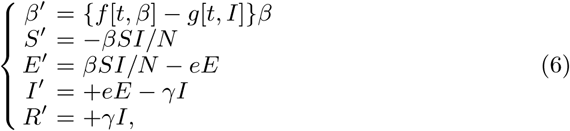

with some non-negative initial conditions *β*(0) = *β*_0_, *S*(0) = *S*_0_, *E*(0) = *E*_0_, *I*(0) = *I*_0_, *R*(0) = *R*_0_, and *f* [*t, β*], *g*[*t, I*] as in Eqns. (5) and (4) respectively. Table 1 describes the variables and parameters of the model.

**Table 1:**
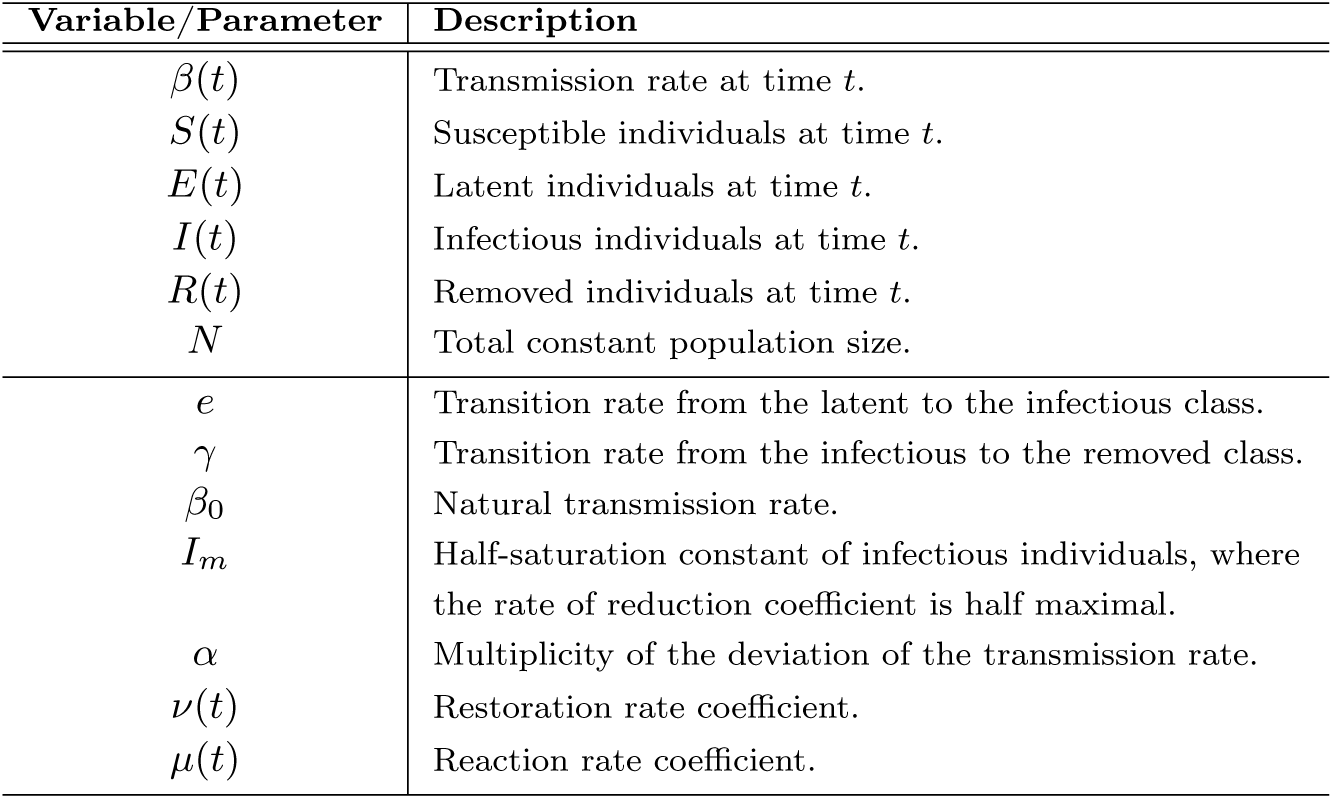
Description of variables and parameters from the model in system (6).

| Variable/Parameter | Description |
| --- | --- |
| $\beta(t)$ | Transmission rate at time $t$. |
| $S(t)$ | Susceptible individuals at time $t$. |
| $E(t)$ | Latent individuals at time $t$. |
| $I(t)$ | Infectious individuals at time $t$. |
| $R(t)$ | Removed individuals at time $t$. |
| $N$ | Total constant population size. |
| $e$ | Transition rate from the latent to the infectious class. |
| $\gamma$ | Transition rate from the infectious to the removed class. |
| $\beta_0$ | Natural transmission rate. |
| $I_m$ | Half-saturation constant of infectious individuals, where the rate of reduction coefficient is half maximal. |
| $\alpha$ | Multiplicity of the deviation of the transmission rate. |
| $\nu(t)$ | Restoration rate coefficient. |
| $\mu(t)$ | Reaction rate coefficient. |

In the following we show that 0 *≤ β*(*t*) *≤ β*_0_, for all *t ≥*0 and *β*_0_ *>* 0. Observe that *β′≥ − µ*(*t*)*β*, and hence due to Grönwall’s inequality (see [38]) we can conclude that 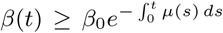. Therefore, *β*(*t*) is non-negative for any non-negative initial condition *β*_0_. We also observe that *β*(*t*) *≡ β*_0_ is an equilibrium solution of the first equation in system (6) as long as *I*(*t*) *≡* 0. In case *I*(0) *>* 0, i.e. disease is present in the population, *β′*(0) *<* 0 and hence *β* (*t*) decreases initially. Since *β′ ≤ν*(*t*) {|*β − β*_0_ |*/β*_0_}^*α*^*β*, using Grönwall’s inequality and the fact that the solutions to the differential equation *β′*= *ν*(*t*) {|*β − β*_0_ |*/β*_0_}^*α*^*β* that pass through points 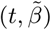 with 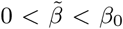 are increasing and bounded by *β*_0_, we obtain that *β*(*t*) *≤ β*_0_ for all *t ≥* 0 and *β*_0_ *>* 0, *I*_0_ *>* 0.

Just as for the classical *SEIR* model, we observe that the epidemiological state variables of the model in system (6) remain positive for positive initial conditions and are bounded by the total population size *N* . Also, adding the second, third and fourth equations in system (6) together we obtain (*S* + *E* + *I*) *′*= *−γI <* 0 whenever *I >* 0. Hence, *S* +*E*+*I* is a non-negative smooth decreasing function, and therefore 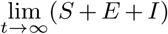 exists. On the other hand, the derivative of any smooth non-negative decreasing function must tend to zero, and hence 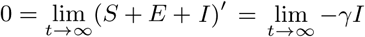, which implies 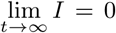. Similarly, by adding the second and third equation in system (6) together, we can prove that 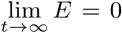. Since the limit of *S* + *E* + *I* exists when time tends to infinity, we can then conclude that 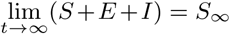. The behavior of *R* can be obtained from *N* = *S* + *E* + *I* + *R* and we obtain 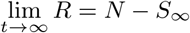. Hence, the long term behavior of the model we present holds (i.e. the limit of the epidemiological state variables exist for infinite time), just as for the classical compartmental epidemic models *SIR* or *SEIR* with constant transmission rate *β* [39]; in particular we have shown that the disease in the long term goes extinct.

Finally, as for classical models we can obtain a threshold condition that determines an initial epidemic outbreak. Given *β*_0_ *>* 0 and considering that *β*(*t*) *≤β*_0_ for all *t ≥*0 as well as that the state variables are bounded by *N*, we have that (*I* + *E*) *′*= *γI* {(*β/γ*)(*S/N*) *−*1} *≤ γI* {(*β*_0_*/γ*) *–* 1} . Hence, we define the basic reproduction number [40, 41] as ℛ_0_ := *β*_0_*/γ*. This way, if ℛ_0_ *<* 1, then the curve representing the infectious population is decreasing to zero and there is no epidemic outbreak. On the other hand, if ℛ_0_ = *β*_0_*/γ >* 1, then (*E* + *I*) increases initially when we assume *S ∼N* and *β*(0) = *β*_0_, and increases as long as (*β*(*t*)*/γ*)*S*(*t*)*/N >* 1 holds. Notice that, in this case, the curve of infectious individuals may be increasing at several time intervals depending on the behavior of the function *β*(*t*), and not only depending on the ratio of susceptible individuals in the population that is decreasing according to the second equation in system (6). In Section 4 we will call the quantity (*β*(*t*)*/γ*)*S*(*t*)*/N* the Effective Reproduction Number, *ℛ*_*e*_, and its dynamics will determine disease dynamics.

In the context of our study, we consider short-medium term scenarios under an epidemic situation, i.e. when ℛ_0_ *>* 1. In particular, we will point out the differences compared to the classical SEIR model with constant *β*, in which *E* and *I* are variables that describe the known unimodal behavior of one bell-shaped curve.

Our model generalizes an idea presented in [42], where the authors consider an SIR-type model with variable transmission rate of the form *β*(*D*(*·*)), in which the time dependent function *D*(*·*) represents social distancing that individuals in the population maintain to each other, governed by the dynamics of *D′*= *−λ*_1_ (*D − D*_*∗*_) + *λ*_2_ *I/N*, where *λ*_1_ and *λ*_2_ are positive constants, and *D*^*∗*^ is the culturally dependent natural social distance, to which *D*(*·*) converges in the absence of disease (*I* = 0). Notice that if in their model 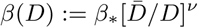 with positive parameters, we observe that 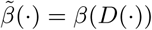 satisfies the equation 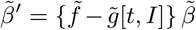, with 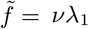 and 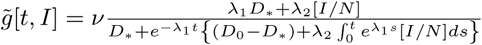 . Observe that regardless of the convergence of 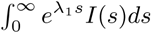, we obtain 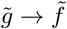 when *t → ∞*.

## 4 Numerical Results

In this section, we present simulations of disease dynamics assuming: first, constant reaction (*µ*(*t*)) and restoration (*ν*(*t*)) coefficients; second, we extend our results to consider time-varying reaction coefficients, representing a diminishing mitigation effect of government responses; third, an additional seasonal effect on the restoration coefficient and we show that our model is capable of capturing real COVID-19 disease data. For the simulations we use the Python programming language [43].

### 4.1 The autonomous *β* SEIR model: constant reaction and restoration coefficients

We consider in this subsection the autonomous *β*SEIR model from system (6), assuming a constant reaction coefficient, *µ*(*t*) = *µ*; i.e. we suppose that the reaction to reduce the transmission rate just depends on point prevalence levels and not on an additional time factor (see Eqn. (4)); and also assuming a constant restoration coefficient, *ν*(*t*) = *ν*; i.e. the regulation on the restoration rate depends only on the deviation of the transmission rate from its natural value *β*_0_ and not on external temporal factors (see Eqn. (5)). We present qualitative results of our model through simulations considering an 18 months time span, and present in each of the Figs. 2-4 five subplots that represent the dynamics for: restoration *f* (*t, β*) and reaction rates *g*(*t, I*); the transmission rate *β*(*t*); the effective reproduction number ℛ_*e*_; the new confirmed cases *eE*(*t*); and finally we illustrate the cumulative number of cases. The effective reproduction number is a time-varying threshold quantity– defined by ℛ_*e*_(*t*) := (*β*(*t*)*/γ*)*S*(*t*)*/N* – such that the number of cases increase while ℛ (*t*) *>* 1, reach a peak when ℛ (*t*) = 1 and decrease when ℛ (*t*) *<* 1 [44, 45], and in particular ℛ_*e*_(0) =ℛ_0_.

**Fig. 2:**
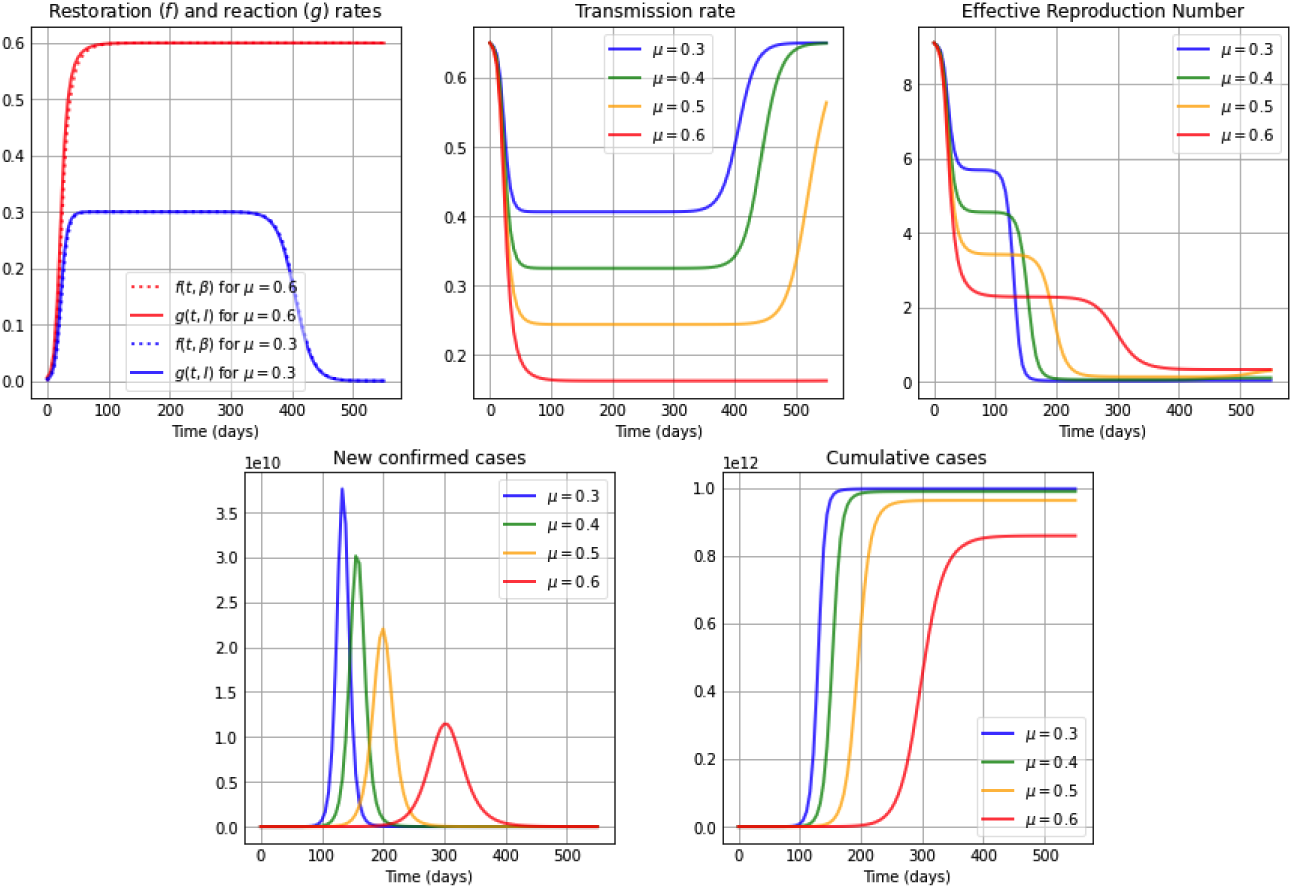
Simulations for a high restoration coefficient *ν* = 0.8. The first subplot illustrates the restoration rate *f* (*t, β*) (dotted) and the reaction rate *g*(*t, I*) (solid). The remaining subplots show: The transmission rate *β*(*t*), the effective reproduction number ℛ_*e*_(*t*), the new confirmed cases *eE*(*t*), and the cumulative cases *E*(*t*) + *I*(*t*) + *R*(*t*). The reaction coefficient in each subplot are chosen to be *µ*: 0.3 (blue); 0.4 (green); 0.5 (orange) and 0.6 (red), and the remaining parameter values and initial conditions are as in Table 2.

Additionally, the constant restoration coefficient *ν* takes in Fig. 2, Fig. 3 and Fig. 4 the values 0.8 (high), 0.5 (medium) and 0.2 (low), respectively, representing high, medium and low rates to restore transmission levels due to the urge to return to the natural transmission rate *β*_0_. For each constant restoration coefficient value, we choose within each figure the constant reaction coefficient *µ* to take the values 0.3 (low, in blue), 0.4 (medium-low, in green), 0.5 (medium-high, in orange) and 0.6 (high, in red), representing different constant levels of the daily maximum reaction to disease that reduces the transmission rate. The remaining parameter values and initial conditions used in the figures are described in Table 2. Note that the basic reproduction number obtained from the values *β*_0_ = 0.65 and *γ* = 1*/*14 from Table 2 is ℛ_0_ = 9.1 *>* 1. For illustration purposes, we use a value significantly larger than one.

**Table 2:** Initial conditions and parameter values and their units, for the simulations in Figs. 2-4 and Figs. 5-7.

| Variable/Parameter | Value | Units |
| --- | --- | --- |
| $\beta_0$ | 0.65 | $(\text{day})^{-1}$ |
| $S_0$ | N | Individuals |
| $E_0$ | 50 | Individuals |
| $I_0$ | 100 | Individuals |
| $R_0$ | 0 | Individuals |
| $N$ | $100^6$ | Individuals |
| $e$ | $1/5$ | $(\text{day})^{-1}$ |
| $\gamma$ | $1/14$ | $(\text{day})^{-1}$ |
| $I_m$ | $10^4$ | Individuals |
| $\alpha$ | 1 | Unitless |

**Fig. 3:**
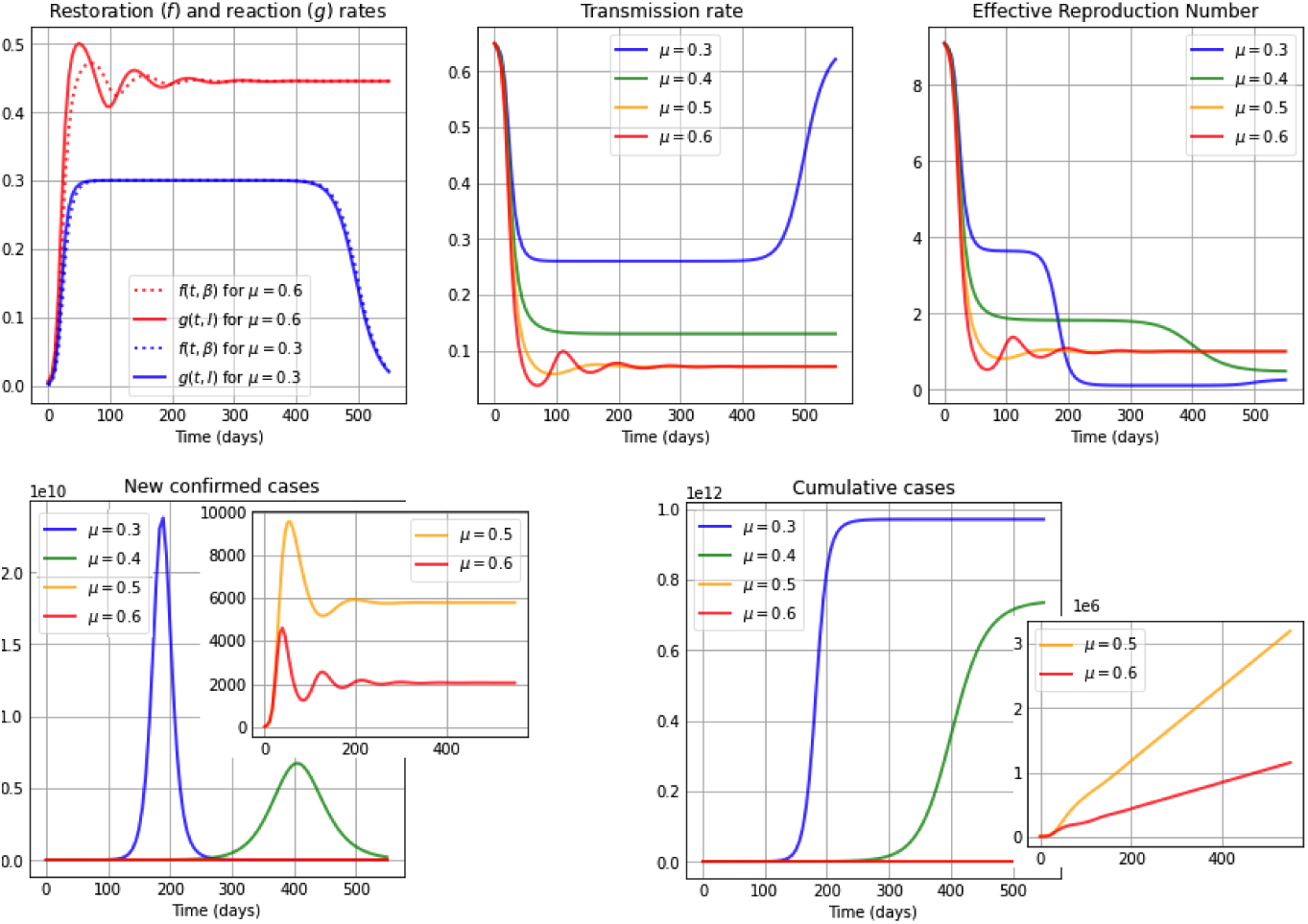
Simulations for a medium restoration coefficient *ν* = 0.5. The first subplot illustrates the restoration rate *f* (*t, β*) (dotted) and the reaction rate *g*(*t, I*) (solid). The remaining subplots show: The transmission rate *β*(*t*), the effective reproduction number ℛ_*e*_(*t*), the new confirmed cases *eE*(*t*), and the cumulative cases *E*(*t*) + *I*(*t*) + *R*(*t*). The reaction coefficient in each subplot are chosen to be *µ*: 0.3 (blue); 0.4 (green); 0.5 (orange) and 0.6 (red), and the remaining parameter values and initial conditions are as in Table 2.

**Fig. 4:**
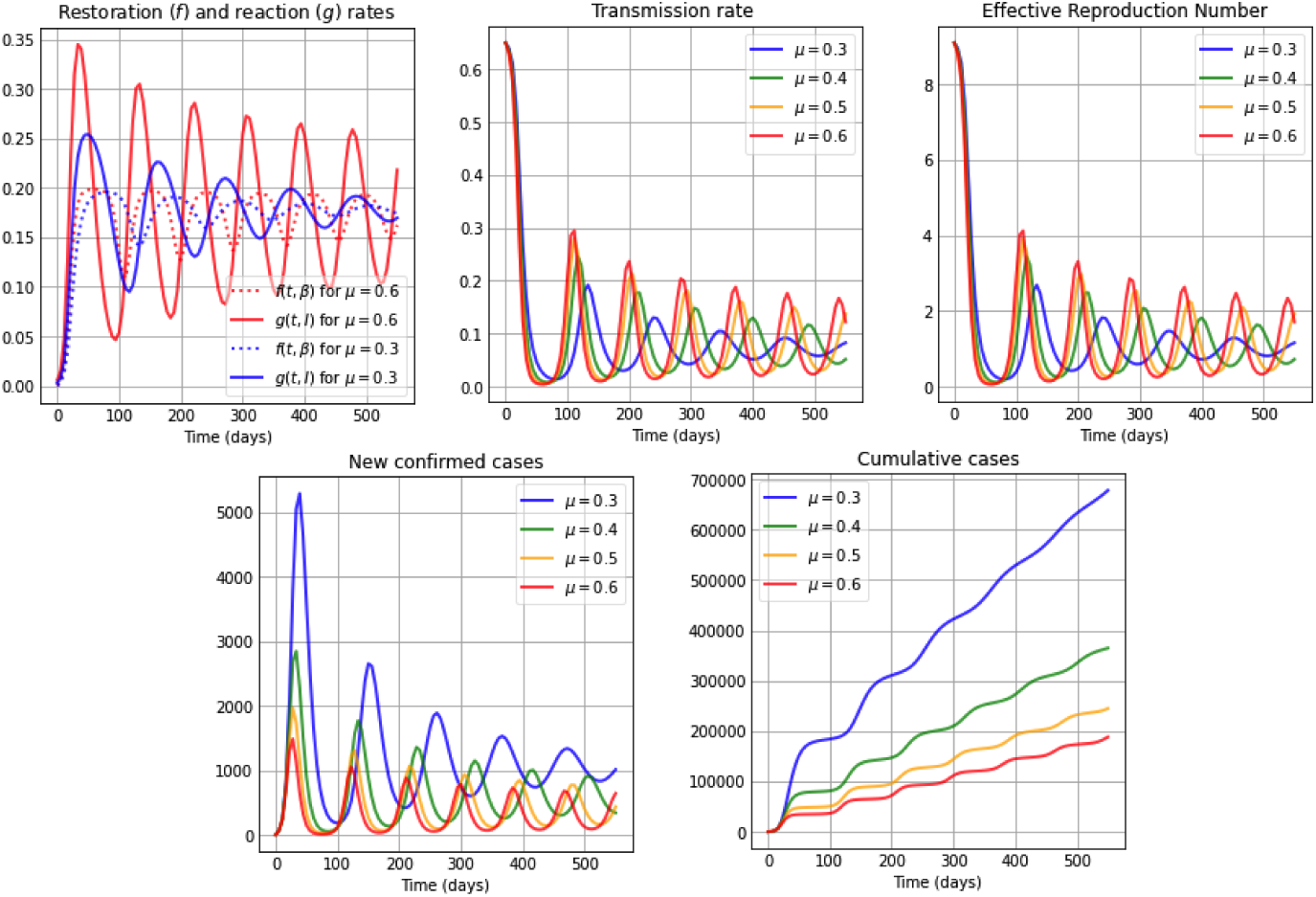
Simulations for a low restoration coefficient *ν* = 0.2. The first subplot illustrates the restoration rate *f* (*t, β*) (dotted) and the reaction rate *g*(*t, I*) (solid). The remaining subplots show: The transmission rate *β*(*t*), the effective reproduction number ℛ_*e*_(*t*), the new confirmed cases *eE*(*t*), and the cumulative cases *E*(*t*) + *I*(*t*) + *R*(*t*). The reaction coefficient in each subplot are chosen to be *µ*: 0.3 (blue); 0.4 (green); 0.5 (orange) and 0.6 (red), and the remaining parameter values and initial conditions are as in Table 2.

Figure 2 considers a high restoration coefficient, *ν* = 0.8. Observe that for high reaction coefficients, e.g. *µ* = 0.6 as well as for small reaction coefficients, e.g. *µ* = 0.3, the restoration rate *f* (*t, β*) and the reaction rate *g*(*t, I*) are very similar to each other. Initially the restoration rate is slightly smaller than the reaction rate, producing that *β* (*t*) *<* 0 (from the first equation in system (6)) and therefore being the reason for the initial decrease of in the transmission rate. After that drop, the reaction and restoration rates are almost equal, producing a plateau in the transmission rate due to *β′* (*t*) *∼*0, and subsequently, a slightly larger restoration rate than reaction rate produces an increase in the transmission rate, which converges to its original natural transmission rate value *β*_0_. Additionally, observe that for all values of the reaction coefficient *µ*, the duration of the plateau in the transmission rate is directly correlated with the value of *µ*: the higher the *µ* value, the longer its duration. Note that, while the transmission rate is at the plateau– i.e. *β*(*·*) behaves similar to constant– when equaling the first equation in our *β*SEIR model (system (6)) to zero, we conclude from *f* (*t, β*) = *g*(*t, I*) when *α* = 1 that

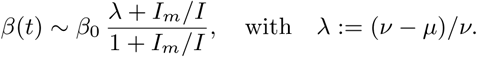

Hence, if *I*_*m*_*≪I*, then *β ∼β*_0_ *λ*. Indeed, notice that the value of *β*_0_ *λ* for *µ* = 0.3, 0.4, 0.5 and 0.6 in Fig. 2 are respectively 0.406, 0.325, 0.244 and 0.163, which correspond very closely to the plateau levels of the respective transmission rates. For all *µ* cases, the effective reproduction number shows a decreasing shape, staying above one at least during the first months of a pandemic. While the effective reproduction number stays above one, one can observe that the number of new confirmed cases increases, reaching a peak when the effective reproduction number reaches the threshold value one. The lower the reaction coefficient value is, the larger is the transmission rate throughout the epidemic, which produces sooner and larger epidemic peaks of new confirmed cases, as well as a rapid increase in the number of cumulative cases, reaching quickly a number close to the final number of infected individuals throughout the whole epidemic. This happens shortly after the effective reproduction number reaches the value of one, and is due to a small number of susceptibles remaining in the population at that time.

Figure 3 considers a medium restoration coefficient, *ν* = 0.5. We observe that for a high reaction coefficient *µ* = 0.6, the restoration (*f* (*t, β*)) and reaction (*g*(*t, I*)) rates show an initial oscillation and differ from each other more clearly, being initially the reaction rate larger than the restoration rate and subsequently both intersecting several times. This produces an oscillatory behavior in the transmission rate due to the *β* equation in system (6), attaining the transmission rate a local maximum or minimum value each time the reaction and restoration rates intersect, i.e. *f* (*t, β*) = *g*(*t, I*). On the contrary, in the case of a low reaction coefficient value, e.g. *µ* = 0.3, we observe very similar restoration and reaction rates (blue curves) just as in Fig. 2. We can also observe from Fig. 3 that a high (red) or medium-high (orange) reaction coefficient *µ*, drives the effective reproduction number below one much faster than in was observed in Fig. 2 (in the case of a higher restoration coefficient) and way before the cases for medium-low (green) and low (blue) reaction coefficient values. It is interesting to see that in the cases of higher reaction coefficients (red and orange), after the initial fast drop of the effective reproduction number, during the remaining time pictured it oscillates around one. When comparing the curves of the new confirmed cases and cumulative cases for these two reaction coefficient values, with the cases of low and medium-low reaction coefficients, we notice that the number of cases is way higher for the latter. Additionally, one bell-shaped curve of new confirmed cases is being observed for smaller reaction coefficient values (blue, green), since the effective reproduction number only manages to cross the threshold ℛ_*e*_ = 1 once due to the small number of susceptibles remaining after that first large peak. On the contrary, an oscillatory behavior is seen for higher *µ* values, obtaining small epidemic peaks each time the effective reproduction number reaches one in a decreasing manner. In other words, the unimodality of the behavior for new confirmed cases observed when *µ* is low (blue) or medium-low (green) is broken for high (red) and medium-high (orange) *µ* values, which was not seen in Fig. 2. One can also observe that for higher *µ* values (red and orange), the increase in the cumulative cases is close to linear in the time-frame pictured, as opposed to the rapid increase in cumulative cases for smaller reaction coefficient values (blue, green).

Figure 4 shows the case of a low restoration coefficient *ν* = 0.2. We observe oscillatory behavior in the restoration and reaction rates, producing an oscillatory behavior in the transmission rate, and hence also in the effective reproduction number and in the number of new confirmed cases. We observe that for high reaction coefficient values, e.g. *µ* = 0.6, the absolute difference between the reaction and restoration rates are larger and their intersections occur sooner, and hence the oscillations in the transmission rate have a higher amplitude and their local maxima occur sooner, than in the case of lower reaction coefficient values, e.g. *µ* = 0.3. Also, transmission rates with higher amplitudes produce less pronounced peaks in the oscillatory behavior of the number of new confirmed cases. Additionally, sooner starting oscillations correspond to higher values of the reaction coefficient *µ*. The cumulative cases show a near to linear increase, where smaller slopes correspond to higher reaction coefficient values.

### 4.2 A non-autonomous *β*SEIR model: time-varying reaction coefficient representing a diminishing mitigation effect

It is to expect that, when a disease enters a population, the reaction coefficient *µ*(*t*) right after the onset increases quickly, reaches a maximum value *µ*_0_, and then decreases due to many factors. This can be deduced (at least) from two sources: (a) The data and information provided by the Oxford COVID-19 Government Response Tracker (OxCGRT) and the time curves defined by the Stringency Index [46, 47]. This index records the intensity of several government responses combined, such as containment and closure policies by country. In general, we observe that, first, the time curves representing the stringency index rise, but then follow a decreasing behavior due to local socioeconomic reasons [48,49, 50, 51, 52,53]. (b) Studies in behavioral science explain the public fall of adherence to mitigation (distancing) measures, as a daily average compliance curve in times of COVID-19 shown for instance in [54] or discussed in the conclusions in [55]. Additionally, it is known that information-based interventions positively impact compliance with mitigation restrictions, such as keeping a certain social distance, decreasing the number of times individuals go out and their time spent outside. Nevertheless, if individuals have been restricted for a prolonged period of time, compliance with such mitigation strategies decreases [56].

We can include such a behavior– representing a diminishing mitigation effect of restrictive measures– through a time-varying reaction coefficient *µ*(*t*), for instance, given by the following Eqn. (7),

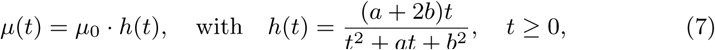

with 0 *< µ*_0_ *<* 1. Notice that *h*(*·*) is a non-negative, unimodal function such that *h′* (*b*) = 0 and *h*(*b*) = 1, i.e. it achieves its maximum value at *t* = *b* and then decreases at a rate that depends on the parameter *a*.

In each of the Figs. 5, 6 and 7 we show for high, medium and low constant restoration coefficients *ν*, respectively, the curves for the restoration (*f* (*t, β*)) and reaction (*g*(*t, I*)) rates, the transmission rate *β*(*t*), the effective reproduction number, the new confirmed cases and the cumulative cases, for a forgetting curve *h*(*t*) of the form given in Eqn. 7, with *a* = 40, *b* = 90, such that the maximum occurs at *t* = 90 days. Within each plot we present curves for different maximum values *µ*_0_ of the now variable reaction coefficient *µ*(*t*) = *µ*_0_*h*(*t*). The other parameters used for these figures are given in Table 2.

**Fig. 5:**
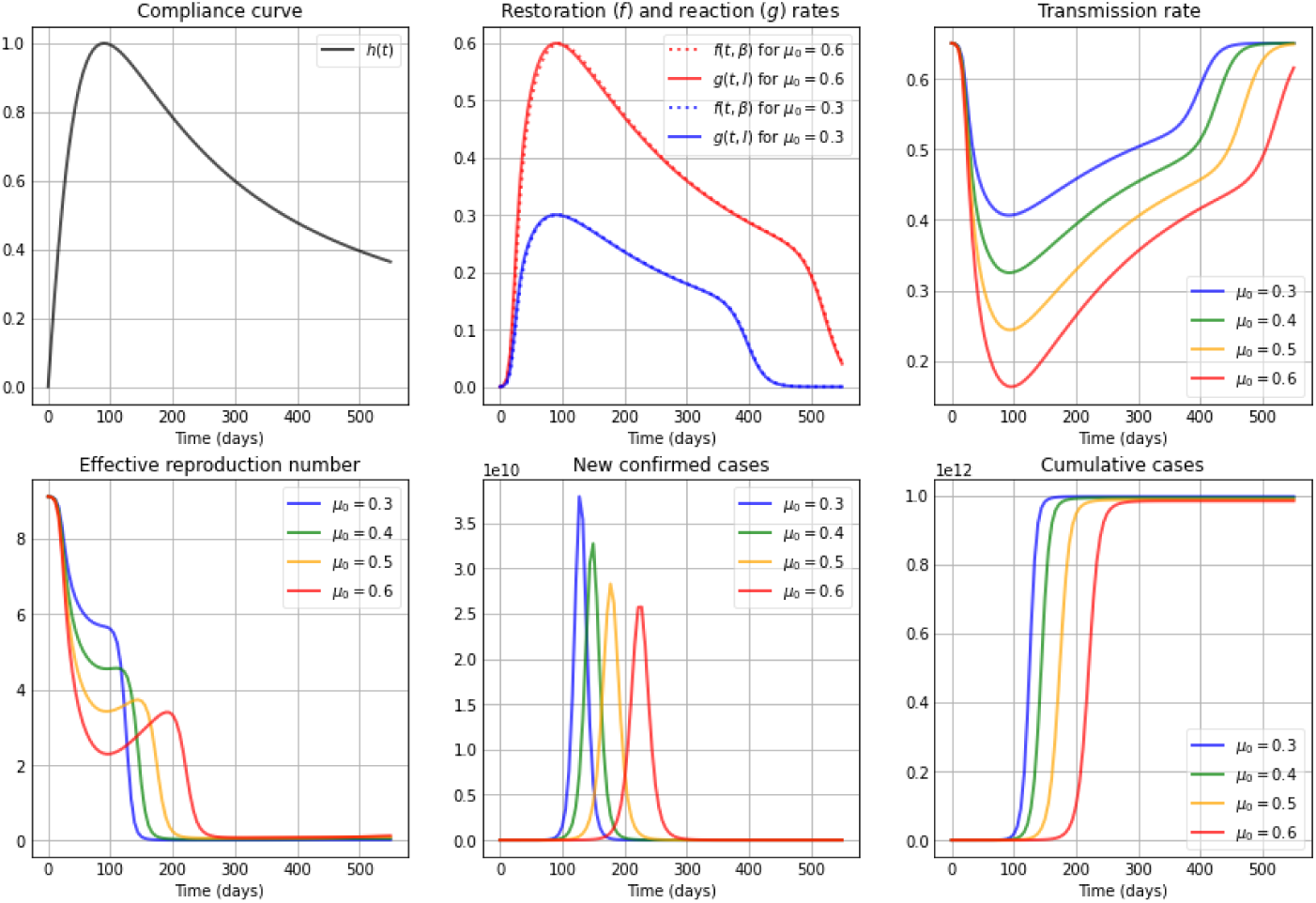
Simulations for a high restoration coefficient *ν* = 0.8. The first subplot depicts the shape of the compliance curve *h*(*t*) = (*a* + 2*b*)*t/*(*t*^2^ + *at* + *b*^2^), with *b* = 90, *a* = 40, and the second plot in the first row illustrates the restoration rate *f* (*t, β*) (dotted) and the reaction rate *g*(*t, I*) (solid). The remaining subplots show: The transmission rate *β*(*t*), the effective reproduction number ℛ_*e*_(*t*), the new confirmed cases *eE*(*t*), and the cumulative cases *E*(*t*) + *I*(*t*) + *R*(*t*). The maximum value *µ*_0_ of the reaction coefficient (*µ*(*t*) = *µ*_0_*h*(*t*)) used in each subplot is: *µ*_0_: 0.3 (blue); 0.4 (green); 0.5 (orange) and 0.6 (red), and the remaining parameter values and initial conditions are as in Table 2.

**Fig. 6:**
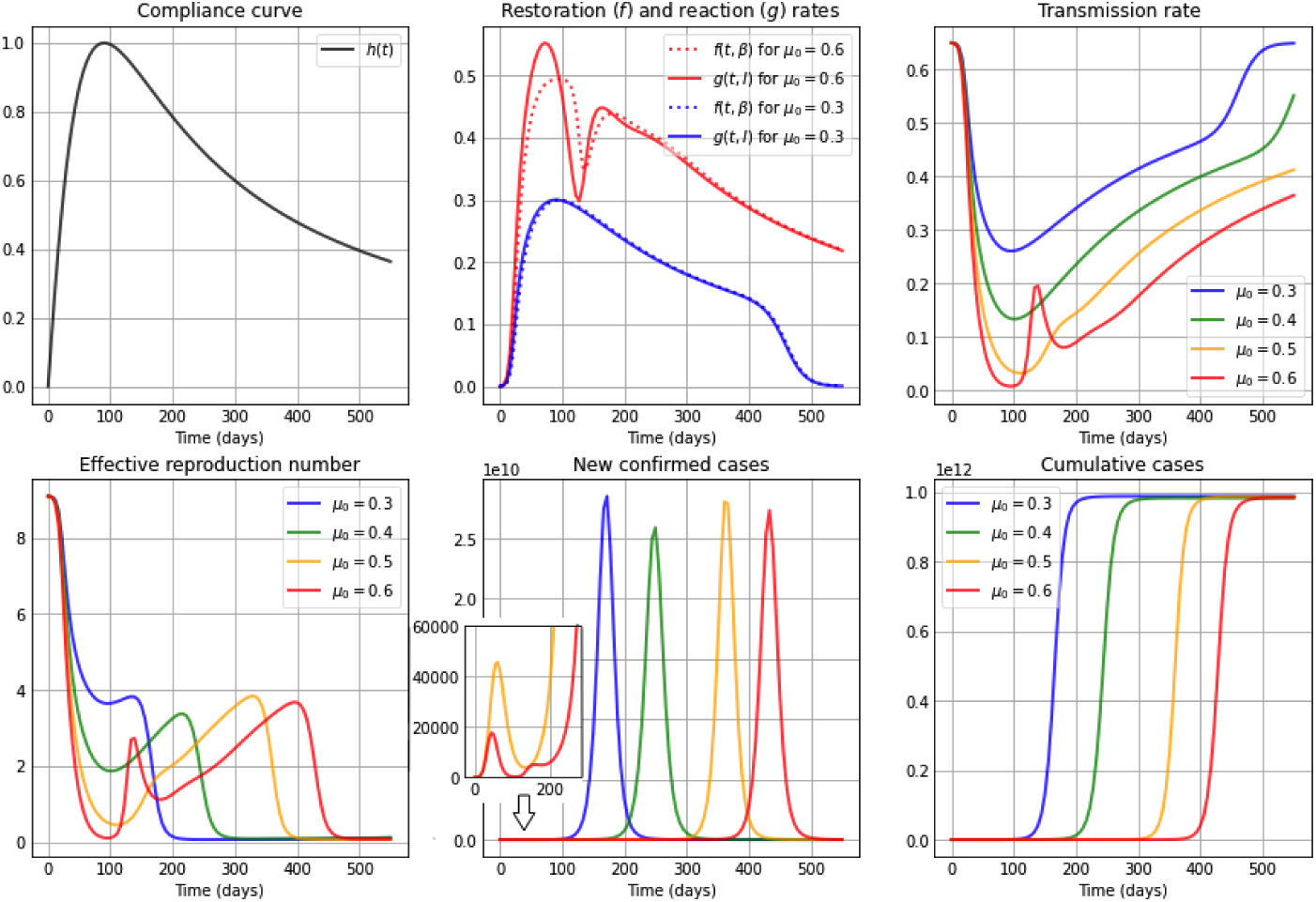
Simulations for a medium restoration coefficient *ν* = 0.5. The first subplot depicts the shape of the compliance curve *h*(*t*) = (*a* + 2*b*)*t/*(*t*^2^ + *at* + *b*^2^), with *b* = 90, *a* = 40, and the second plot in the first row illustrates the restoration rate *f* (*t, β*) (dotted) and the reaction rate *g*(*t, I*) (solid). The remaining subplots show: The transmission rate *β*(*t*), the effective reproduction number ℛ_*e*_(*t*), the new confirmed cases *eE*(*t*), and the cumulative cases *E*(*t*) + *I*(*t*) + *R*(*t*). The maximum value *µ*_0_ of the reaction coefficient (*µ*(*t*) = *µ*_0_*h*(*t*)) used in each subplot is: *µ*_0_: 0.3 (blue); 0.4 (green); 0.5 (orange) and 0.6 (red), and the remaining parameter values and initial conditions are as in Table 2.

**Fig. 7:**
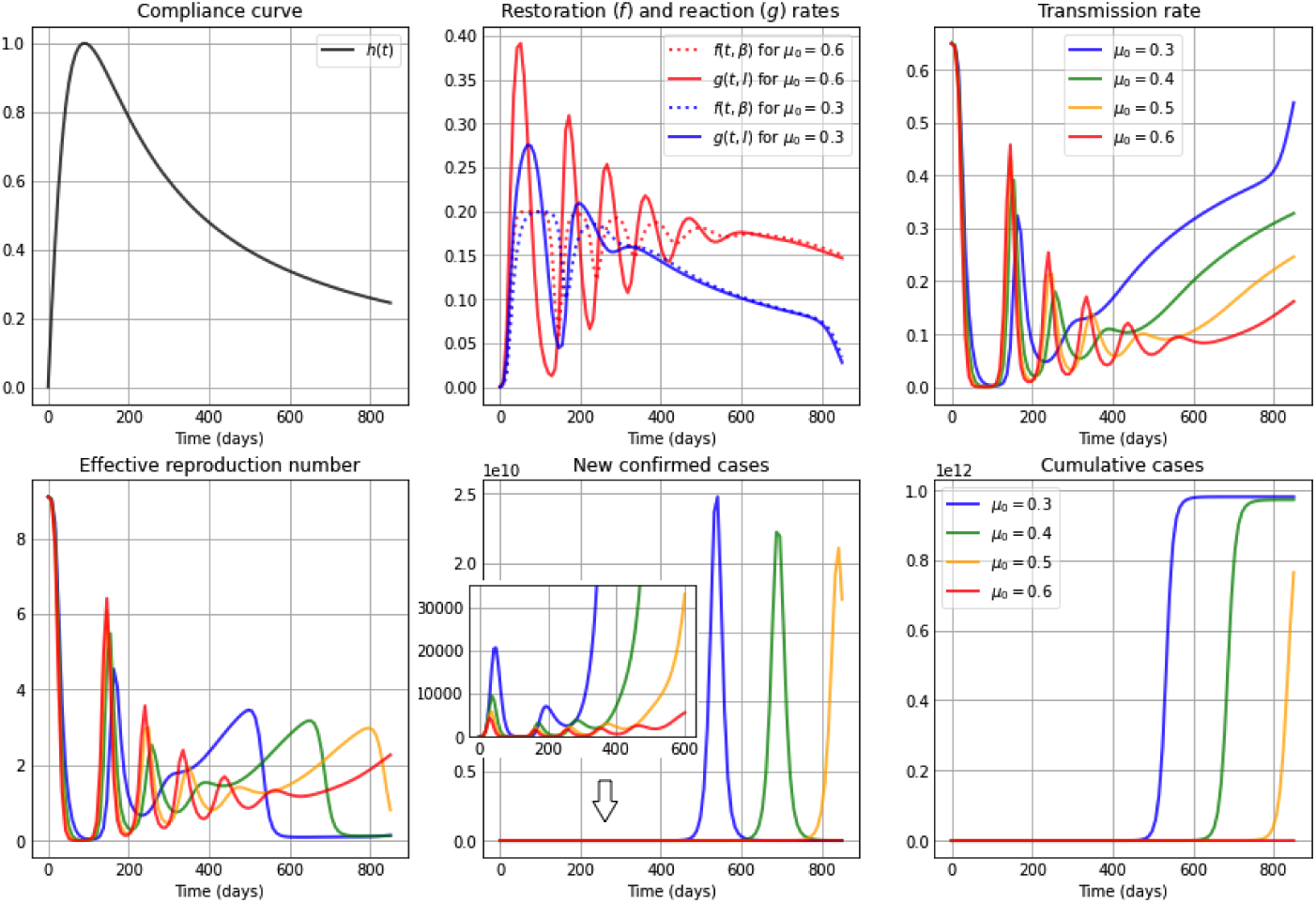
Simulations for a low restoration coefficient *ν* = 0.2. The first subplot depicts the shape of the compliance curve *h*(*t*) = (*a* + 2*b*)*t/*(*t*^2^ + *at* + *b*^2^), with *b* = 90, *a* = 40, and the second plot in the first row illustrates the restoration rate *f* (*t, β*) (dotted) and the reaction rate *g*(*t, I*) (solid). The remaining subplots show: The transmission rate *β*(*t*), the effective reproduction number ℛ_*e*_(*t*), the new confirmed cases *eE*(*t*), and the cumulative cases *E*(*t*) + *I*(*t*) + *R*(*t*). The maximum value *µ*_0_ of the reaction coefficient (*µ*(*t*) = *µ*_0_*h*(*t*)) used in each subplot is: *µ*_0_: 0.3 (blue); 0.4 (green); 0.5 (orange) and 0.6 (red), and the remaining parameter values and initial conditions are as in Table 2.

We present in Fig. 5 the case of a high constant restoration coefficient *ν* = 0.8. We observe that the restoration and reaction rates are very similar regardless of the *µ*_0_ value. Despite their similarity, initially, up to day 90, a slightly higher reaction than restoration rate produces a sharp decrease in the transmission rate. The decreasing shape of the forgetting curve starting on day 90, immediately reduces the reaction rate to slightly below the restoration rate, producing an increase in the transmission rate, eliminating the plateau observed in Fig. 2, in which the reaction coefficient was assumed constant. Due to the incorporation of a forgetting-curve in the reaction coefficient we can also observe higher and earlier occurring epidemic peaks in the curves of new confirmed cases as compared to the constant reaction coefficient case (see Fig. 2), especially for large *µ*_0_ values.

Figure 6 depicts the dynamics for a medium restoration coefficient *ν* = 0.5. Here we can observe– especially for a large maximum value *µ*_0_ of the reaction coefficient *µ*(*t*) = *µ*_0_*h*(*t*)– that the restoration rate *f* (*t, β*) and the reaction rate *g*(*t, I*) differ more from each other, as compared to the case depicted in Fig. 5. If we observe the curves for *µ*_0_ = 0.6 we see that initially, up to day 90, the reaction rate is clearly higher than the restoration rate, which again produces a sharp decrease in the transmission rate. Every time the restoration and reaction rate curves intersect, we can observe a local maximum/minimum in the transmission rate, which eventually increases and converges back to its natural value *β*_0_. The changing human behavior reflected in the restoration and reaction rates, observed for *µ*_0_ = 0.6, produces a late occurring but large epidemic peak, preceded by one small peak. This dynamics can also be explained by observing the shape of the effective reproduction number, which crosses the threshold ℛ_*e*_ = 1 three times (red curve).

Figure 7 shows the dynamics for a low constant restoration coefficient *ν* = 0.2. Here we observe initial oscillatory dynamics for the restoration and reaction rates, which explain the oscillatory dynamics in the transmission rate and also in the effective reproduction number, which crosses the threshold ℛ_*e*_ = 1 several times, producing small oscillations in the new confirmed cases, followed by a large epidemic peak.

### 4.3 A non-autonomous *β* SEIR model and the example of COVID-19 in Chile and Italy: time-varying reaction and restoration coefficients

Although it has not been proven that the virus SARS-CoV-2 is seasonal in nature, seasonality of transmission may be an important factor to consider since social behavior is environmentally driven [57]. The main hypothesis regarding seasonality indicates that the higher the temperature the fewer infections occur [58, 59]. Therefore, additional to a time-varying reaction rate, the restoration rate could depend, among other, on environmental factors such as for instance temperature. For example, in regions with Mediterranean climate, an annual oscillation of the daily mean temperature can be observed [60], as is the case in a large part of Italy or central Chile. The seasonality could also represent peoples’ behavior during winter months vs summer months, or during vacation periods. This affects the tendency of the population to return to its natural transmission rate *β*_0_, while making it seasonal in nature. Hence, to capture seasonal factors we consider an annual periodic restoration coefficient of the form

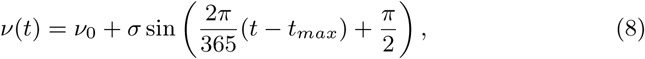

where *ν*_0_ is the average restoration coefficient value, *σ* the amplitude of the oscillation and *t*_*max*_ the moment in time, where the restoration coefficient is maximum.

Considering this non-constant seasonal behavior for the restoration coefficient *ν*(*t*) as given in Eqn. (8), as well as a time-varying reaction coefficient as described in Eqn. (7), we illustrate through Fig. 8 and Fig. 9 the applicability of our model during the first months of the pandemic, by performing a simple fit to COVID-19 data of new confirmed cases of Chile and Italy [61, 62], respectively, which uses the *minimize()* function and the *leastsq* method in Python. We use Chile and Italy as examples of countries at different hemispheres that experienced COVID-19 in distinct ways, due to cultural differences, distinct levels of initial knowledge of the virus, seasons, etc., and this way to show that our model can capture different COVID-19 dynamics and partially describe them through reaction and restoration thrives of the population. Table 3 and Table 4 show the respective fixed and fitted parameter values used in each figure.

**Table 3:** Parameters for the simulations in Fig. 8 for the case of Chile.

| Fixed parameters | Value | Reference |
| --- | --- | --- |
| $e$ | 1/5 | [63, 64] |
| $\gamma$ | 1/14 | [65, 66] |
| $\mathcal{R}_0$ | 5.4 | [12] |
| $\beta_0$ | 0.38 | Assumed as $\mathcal{R}_0\gamma$ |
| $b$ | 110 | Adapted from [46, 47] |
| $a$ | 500 | Adapted from [46, 47] |
| $I_m$ | 7200 | Assumed |
| $\alpha$ | 1 | Assumed |
| Fitted parameters | Fitted Value |  |
| $\mu_0$ | $0.79406527 \pm 0.00241983$ | |
| $\nu_0$ | $0.76149013 \pm 0.00663077$ | |
| $\sigma_0$ | $0.14000051 \pm 0.00641483$ | |
| $t_{max}$ | $41.2846863 \pm 1.96538852$ | |

**Table 4:** Parameters for the simulations in Fig. 9 for the case of Italy.

| Fixed parameters | Value | Reference |
| --- | --- | --- |
| $e$ | 1/5 | [63, 64] |
| $\gamma$ | 1/14 | [65, 66] |
| $\mathcal{R}_0$ | 5.8 | [67, 68, 69] |
| $\beta_0$ | 0.414 | Assumed as $\mathcal{R}_0\gamma$ |
| $b$ | 67 | Adapted from [46, 47] |
| $a$ | 80 | Adapted from [46, 47] |
| $I_m$ | 12100 | Assumed |
| $\alpha$ | 1 | Assumed |
| Fitted parameters | Fitted Value |  |
| $\mu_0$ | $0.76946067 \pm 0.00138200$ | |
| $\nu_0$ | $0.66441900 \pm 0.00645309$ | |
| $\sigma_0$ | $0.49996423 \pm 0.00936939$ | |
| $t_{max}$ | $315.955940 \pm 0.62948680$ | |

**Fig. 8:**
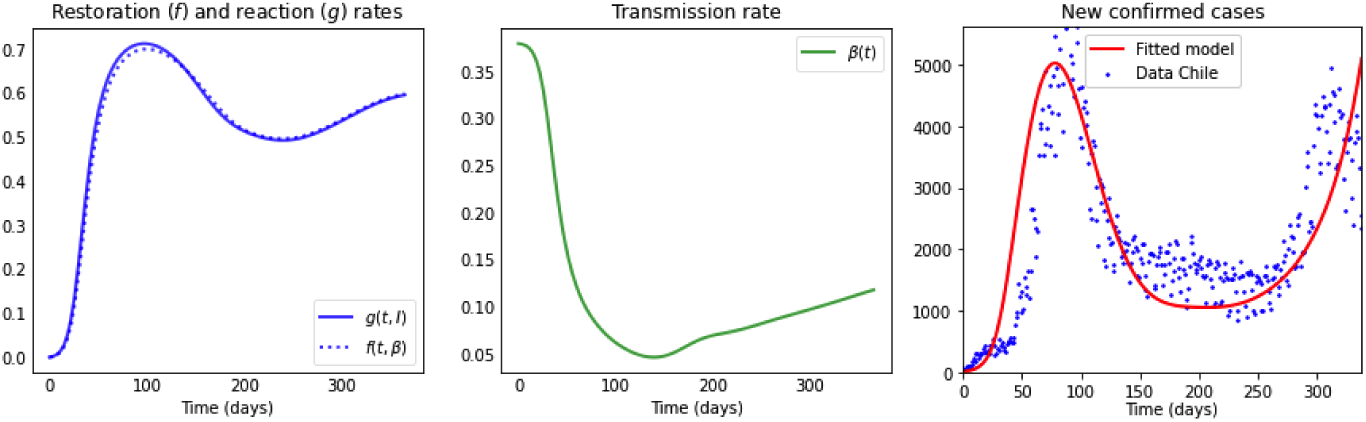
The first subplot depicts the restoration rate *f* (*t, β*) (dotted) and reaction rate *g*(*t, I*) (solid); the second plot the transmission rate *β*(*t*); and in the third plot the blue dots represent data of daily confirmed new cases of COVID-19 in Chile, from March 16th, 2020, to February 16th, 2021 [61]. The red curve represents the least square fit of the model to the data with parameter values as in Table 3 for the population of Chile, with *N* = 18 million individuals and initial conditions of the model (6) taken to be *E*_0_ = 20, *I*_0_ = 81, *R*_0_ = 0, *S*_0_ = *N − E*_0_ *− I*_0_ *− R*_0_.

**Fig. 9:**
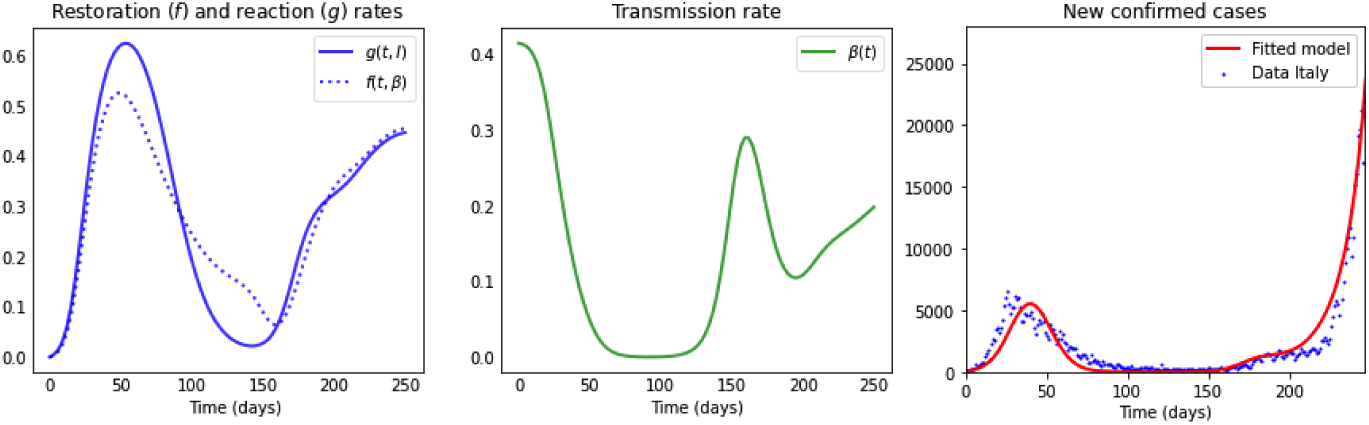
The first subplot depicts the restoration rate *f* (*t, β*) (dotted) and reaction rate *g*(*t, I*) (solid); the second plot the transmission rate *β*(*t*); and in the third plot the blue dots represent data of daily confirmed new cases of COVID-19 in Italy, from February 24th, 2020, to October 31st, 2020 [62]. The red curve represents the least square fit of the model to the data with parameter values as in Table 4 for the population of Italy, with *N* = 60.5 million individuals and initial conditions of the model (6) taken to be *E*_0_ = 81, *I*_0_ = 566, *R*_0_ = 0, *S*_0_ = *N − E*_0_ *− I*_0_ *− R*_0_.

We can observe how the restoration rate and reaction rate differ among countries: In the case of Chile (see Fig. 8), they are very similar, expecting a population whose reaction to disease presence and urge to return to their normal behavior (restoration) is similar in nature. Nevertheless, a small difference in those rates produce a large impact on the transmission rate, such as the steep decrease observed initially.

On the other hand, from Fig. 9 one can see that the reaction rate (solid curve) and the restoration rate (dotted curve) differ more than in the case of Chile, having initially a population, which is reacting to disease presence faster than their urge to restore normal conditions, for later reversing their behavior three times, producing during that time period a long lasting plateau. Finally, one can see that a larger restoration rate than reaction rate produces the appearance of a large second peak.

## 5 Discussion and Conclusions

In [70](1973) and [71] (1989), strategic models are defined as those that, despite not having high resolution concerning a specific reality, have the advantage of containing all the minimum aspects of the referenced system. The main contribution of this work is to achieve a dynamically richer low-cost model, that is, one that adds only one more differential law to the classical SEIR model (without introducing new compartments in the population).

There is evidence in the literature that populations change their behavior when facing dangerous diseases, i.e., reacting to these by managing to modify the transmission rate. Our proposed model, the *β*SEIR model, is a serious candidate to contain the minimum aspects for disease transmission of a high impact infectious-contagious disease in populations that, while living with the urge to restore normal conditions, react to reduce favorable conditions for disease transmission. Specifically, the novelty of the *β*SEIR model we present is, that it incorporates a variation in the transmission rate, which occurs proportional to the transmission rate itself but also proportional to a tension given by the difference (*f − g*) between a) reaction rate (*g*) to disease presence that may include other behavioral factors, such as compliance with mitigation strategies, and b) restoration rate (*f*) that aims to restore a certain intrinsic value of disease transmission, due to for instance socio-environmental elements.

Our results show an important gain in dynamic possibilities even in the case where Eqn. (3) in the *β*SEIR model (6) is autonomous, i.e. *f* and *g* do not depend explicitly on time. Indeed, we can see in Figs. 2-4 the appearance of several epidemic peaks and initial oscillatory dynamics, explained by the tension between reaction and restoration thrives of a population. In particular, we observe that high restoration coefficients *ν*– affecting the restoration rate *f* and representing a higher urge of the population to return to normal conditions– induce temporary stabilization of the transmission rate after an initial drop, being the duration of this plateau larger, the larger the reaction coefficients *µ* (affecting the reaction rate *g*) are, i.e. the higher the self-protective reaction is to disease presence. When considering small *ν* values (a small urge to return to normality), we observe oscillations in the transmission rate, with higher amplitudes for higher *µ* values, i.e. amplitudes are higher if individuals’ reaction to disease presence is higher. These oscillations in the transmission rate generate oscillations in the effective reproduction number, which lay around one for a period of time proportional to the value of the reaction coefficient *µ*. Regarding the curve of new cases, we show that in general higher restoration coefficients *ν* produce unimodal behavior, whereas lower *ν* values generate the appearance of a finite number of peaks with decreasing peak size, in a way that for large reaction coefficients *µ* the timing between peaks is smaller. These results already define interesting future mathematical challenges.

In general, our results show how the transmission rate is impacted by the reaction rate *g* and the restoration rate *f* . In particular, we observe that a small difference between reaction and restoration rates may produce a large impact in the transmission rate. This highlights the importance of individual behavior in a pandemic setting, where even the behavior of a small number of individuals could change the dynamics of a disease drastically.

The real data of new confirmed cases presented in Fig. 1 show curves with two and up to three waves, differing among countries in time and size, with peaks and valleys of different heights, proper to a pandemic still under development. We notice that the *β*SEIR model can capture such patterns at the cost of varying the reaction coefficient *µ*(*t*) and for further dynamic richness varying the restoration coefficient *ν*(*t*). We can justify a time-varying reaction coefficient *µ*(*t*) that considers two aspects: first, it follows the shape of the Stringency-Index [46] – that records for each country government mitigation measures–, second, it reflects the reduced compliance with or adherence to mitigation strategies observed with time. The *β*SEIR model shows even richer dynamics when introducing such a time-varying reaction coefficient (see Figs. 5-7).

Additionally, our model is capable of capturing the time series of new confirmed cases of Chile and Italy when including– on top of a time-varying reaction coefficient– a time-dependent seasonal variation in the restoration coefficient *ν*(*t*), reflecting distinct temporal and possibly behavioral characteristics of two countries located at different hemispheres. These results are, at first glance, good indicators for the richness that a model of such low structural complexity, as the one proposed here, can provide.

## Data Availability

We used publicly available data of new confirmed cases of Chile and Italy from the links below.

https://www.minciencia.gob.cl/covid19/

https://www.statista.com/statistics/1101690/coronavirus-new-cases-development-italy/

